# Deep Learning Decodes Latent ECG Signatures of Stress Cardiomyopathy

**DOI:** 10.64898/2026.01.21.26344596

**Authors:** Jooyoung Ryu, Carl Harris, Chenjia Zhang, Kirby Gong, Robert D. Stevens

## Abstract

**Background:** Stress cardiomyopathy (SCM) shares features with acute myocardial infarction (AMI) which may lead to misdiagnosis and misaligned management decisions. We hypothesize that features derived from the 12-lead ECG can identify cases of SCM and differentiate them from AMI.

**Methods:** Using data from a large registry of critically ill patients, we trained a deep learning algorithm to perform two classification tasks: (1) SCM vs. Non-SCM and (2) SCM vs. AMI. Model training was accomplished with 3 different sets of input features: raw ECG waveforms, clinical features from the electronic health record (EHR), and a fusion model combining ECG with clinical data. SHapley Additive exPlanations (SHAP) analysis was performed to identify the most influential predictive features.

**Results:** Among 71,479 patients admitted to ICU, 349 (0.48%) were diagnosed with SCM while 4,507 (6.31%) had AMI. The clinical-ECG fusion model achieved best performance, with area under the precision-recall curve (AUPRC) values of 0.191 (0.146-0.243) for SCM vs. Non-SCM and 0.430 (0.371-0.488) for SCM vs. AMI, outperforming baseline AUPRCs of 0.0048 and 0.0625, respectively. The fusion model outperformed both the waveform model (p < 0.001) and the EHR model (p < 0.001) for both SCM vs. Non-SCM and SCM vs. AMI. The waveform model achieved AUPRC values of 0.089 (0.062-0.125) for SCM vs. Non-SCM and 0.309 (0.257-0.374) for SCM vs.

AMI. There was no statistically significant difference in performance between the waveform model and the EHR model for both tasks. SHAP analysis highlighted female gender as well as congestive heart failure (CHF) and hypertension as the features most influential in predicting SCM.

**Conclusion:** Findings indicate that ECG waveforms contain latent information which supports the detection of SCM in patients admitted to the ICU. ECG-based deep learning screening could enable early identification and treatment of SCM and might be particularly valuable in resource-constrained environments.

## Introduction

Stress cardiomyopathy (SCM), also known as Takotsubo syndrome or broken heart syndrome, is a type of acute cardiac dysfunction triggered by physical or psychological stress ^1,2^. While SCM risk factors have been identified and an association with increased adrenergic signaling has been reported, the biological mechanisms driving SCM remain poorly understood ^1,3^. SCM is a serious life-threatening condition that carries a significant risk of death and major adverse cardiovascular events (MACE)^3^. Furthermore, the clinical presentation of SCM may closely resemble acute myocardial infarction (AMI) ^3,5^, yet these two conditions require radically different treatments. It follows that early and accurate differentiation between SCM and AMI is a clinical priority ^8,9^.

Although diagnostic criteria for SCM have been proposed, there is a lack of alignment on the optimal diagnostic approach ^10^. Clinicians generally rely on a combination of medical history, ECG, echocardiography, and troponin levels to confirm the diagnosis ^15^. In equivocal cases, patients may undergo coronary angiography^11,12^ or cardiac MRI^13–16^. The latter tests pose risks to patients, are resource-intensive, time-consuming, and require specialized expertise, making them unsuitable for low-resource settings or situations that require rapid decision-making ^14^.

The 12-lead electrocardiogram (ECG) is low-cost and widely accessible. While previous studies have identified ECG abnormalities associated with SCM, such as ST-segment elevation, T-wave inversion, and QT-interval prolongation, these studies are hard to generalize due to limited sample sizes and the ECG changes identified are not well defined or require expert interpretation ^17–21^. Moreover, previous computational SCM studies relied on ECG-derived numerical parameters rather than waveforms ^17,22^. The extraction of these parameters can be labor-intensive, error-prone, and inconsistent across different ECG machines and hospitals ^23,24^.

The primary aim of this study is to identify SCM patients and differentiate them from AMI by applying a deep learning analytical pipeline to raw ECG data. We hypothesize that SCM has distinct ECG signatures which can be effectively decoded using deep learning. Deep learning has been used to analyze ECG signals directly for identifying various cardiac diseases, bypassing the need for manual feature extraction or preprocessing ^24^. This captures subtle, nuanced ECG patterns, potentially increasing diagnostic accuracy.

## Methods

### Approach

This study focuses on two binary classification tasks: distinguishing SCM from Non-SCM and differentiating SCM from AMI. For each task, three deep learning models were developed: an ECG-only waveform model that uses only ECG waveforms as input, an EHR model with only clinical features, and a fusion model that combines the ECG waveforms with clinical features.

### Data Sources

The data source was the Medical Information Mart for Intensive Care IV (MIMIC-IV) database, a publicly available clinical dataset from the PhysioNet repository ^25,26^. The MIMIC-IV database is a comprehensive collection of de-identified electronic health data of patients admitted to the Beth Israel Deaconess Medical Center in Boston, Massachusetts.

Specifically, we utilized three main modules within MIMIC-IV for patient selection and feature extraction. The MIMIC-IV hosp module contains electronic health record (EHR) such as demographic information, laboratory results, and International Classification of Diseases (ICD) diagnosis codes which served as ground truth labels for patient selection. The MIMIC-IV-ECG module contains approximately 800,000 ECG records from nearly 160,000 unique patients. This dataset provided access to raw 12-lead diagnostic ECG waveforms, essential for identifying cardiac conditions such as SCM and AMI. Lastly, the MIMIC-IV-Note module includes de-identified clinical notes, providing detailed discharge summaries including medical history and ongoing medical conditions.

### Selection Criteria

Figure 1 illustrates the patient selection process. A total of 299,712 adult patients (≥18 years old) were identified in the MIMIC-IV dataset. Patients were excluded if they met any of the following criteria: no hospital admission record, no discharge notes, no ECG records, ECG with missing data, or ECG records taken outside the hospital admission period. The resulting sample of 71,479 patients was then divided into 349 (0.48%) SCM patients and 71,130 (99.51%) non-SCM patients using ICD-9/10 codes. The non-SCM cohort was further divided into 4,507 (6.31%) AMI patients and 66,623 (93.21%) non-AMI patients.

**Figure 1.**
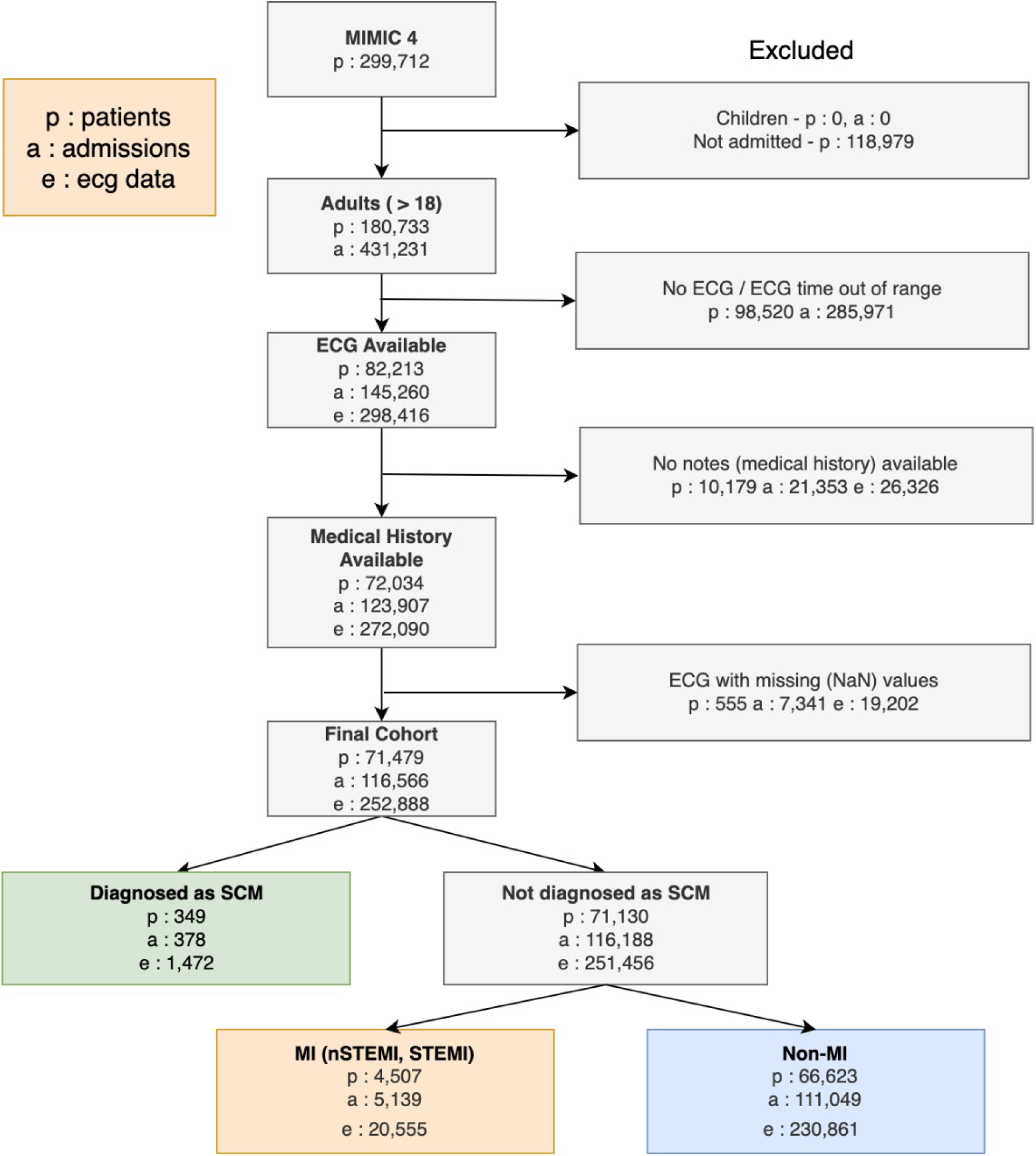
**Flowchart of patient selection process**

Patients were assigned exclusively to one group to prevent data leakage. Patients diagnosed with SCM were included only in the SCM group and removed from the non-SCM group, even if they had additional diagnoses including AMI. The non-SCM group was similarly divided into AMI and non-AMI cohorts. For the SCM vs. AMI task, patients with dual diagnoses of SCM and AMI (n = 62) were excluded to avoid label ambiguity.

### Cohort Identification and Labeling Strategy

ICD-9/10 codes were utilized as ground truth labels. SCM was labeled as 1 and non-SCM was labeled as 0 for SCM vs. non-SCM. For the SCM vs. AMI task, we restricted the cohort to admissions labeled as SCM or AMI only. SCM was labeled as 1 and AMI as 0. The specific ICD codes used to identify SCM and AMI are listed in Supplementary Table 1.

ICD codes within the MIMIC-IV dataset were assigned on the admission level, indicating all diagnostic conditions and diseases the patient had during each admission period. Multiple ICD codes were often assigned to a single admission, as it is common for a patient to have multiple conditions and diagnoses during admission. Admissions were labeled as SCM if any ICD code within the admission equaled ICD codes specific for SCM (‘I51.81’ for ICD-10 and ‘42983’ for ICD-9). Similarly, within the non-SCM cohort, admissions were identified as AMI if any ICD code within the admission equaled ICD codes specific for AMI (listed in Supplementary Table 1). Admissions without ICD codes for either SCM or AMI were identified as non-AMI cohort. All codes were validated by a fellowship-trained, board-certified intensive care physician.

### Feature Extraction

All feature extraction and preprocessing steps were reviewed by a fellowship-trained, board-certified intensive care physician to ensure clinical relevance and accuracy of extracted feature values. The same feature extraction and preprocessing was applied across all models.

Raw 12-lead diagnostic ECG waveforms were extracted from the MIMIC-IV ECG subset. Each waveform was 10 seconds in length, recorded at a sampling rate of 500 Hz. These waveforms were used as inputs for both the waveform and fusion model.

Predictive variables for the fusion and EHR model were selected through clinician input and literature review. A total of 40 clinical features were identified, including demographics, cardiac biomarkers, comorbidities, and medication administration. Comorbidities were identified using the Elixhauser Comorbidity Index conversion table described by Quan et al ^27^.

A total of seven additional binary features were created, five for medication administration and two for past medical history. Medication features focused on four catecholamines (dopamine, dobutamine, epinephrine, norepinephrine), which are neurotransmitters and medications commonly used to treat conditions such as cardiogenic shock, cardiac dysfunction, anesthesia, and anaphylaxis. Catecholamines administered for anesthesia and anaphylaxis were excluded from the extraction process. All administration of these four catecholamines were recorded as continuous IV infusions. For each catecholamine, a binary variable was set to 1 if the drug was administered during the admission period. An additional binary variable called “catecholamine” was set to 1 if any of the four drugs were administered.

Two binary features were extracted for history of psychological or neurologic disorders, which were identified as key risk factors of SCM according to the InterTak Diagnostic Score, a clinical tool designed to differentiate SCM from other cardiac conditions based on patient characteristics and risk factors ^28^. These features were derived from the “Past Medical History” section of each admission’s discharge notes using keyword matching, where a value of 1 indicates that at least one of the corresponding keywords appeared in the discharge report.

A full list of clinical features, comorbidities, excluded catecholamine drugs, and keywords used for medical history extraction is provided in Supplementary Table 2.

### Preprocessing

Age was standardized using z-score normalization. No additional preprocessing was applied to ECG waveforms, as ECGs with missing values were already excluded during patient selection.

We randomly split data between training, validation, and test sets on unique patient ID to prevent data leakage (i.e., such that the same patient would not appear in both the training and test sets). During training, all ECGs within admission were used to allow the model to learn from more samples, especially considering the small number of positive SCM samples. For the test set, however, a 24-hour filter was applied to test the model’s ability to predict SCM early within admission. For a single admission, only the first ECG record within 24 hours was used. If there was no ECG record within 24 hours upon admission, that admission was removed from the test set.

### Models

Figure 2 provides a schematic overview of the modeling approach described in this section, illustrating key differences in fusion architecture and modeling strategies.

**Figure 2.**
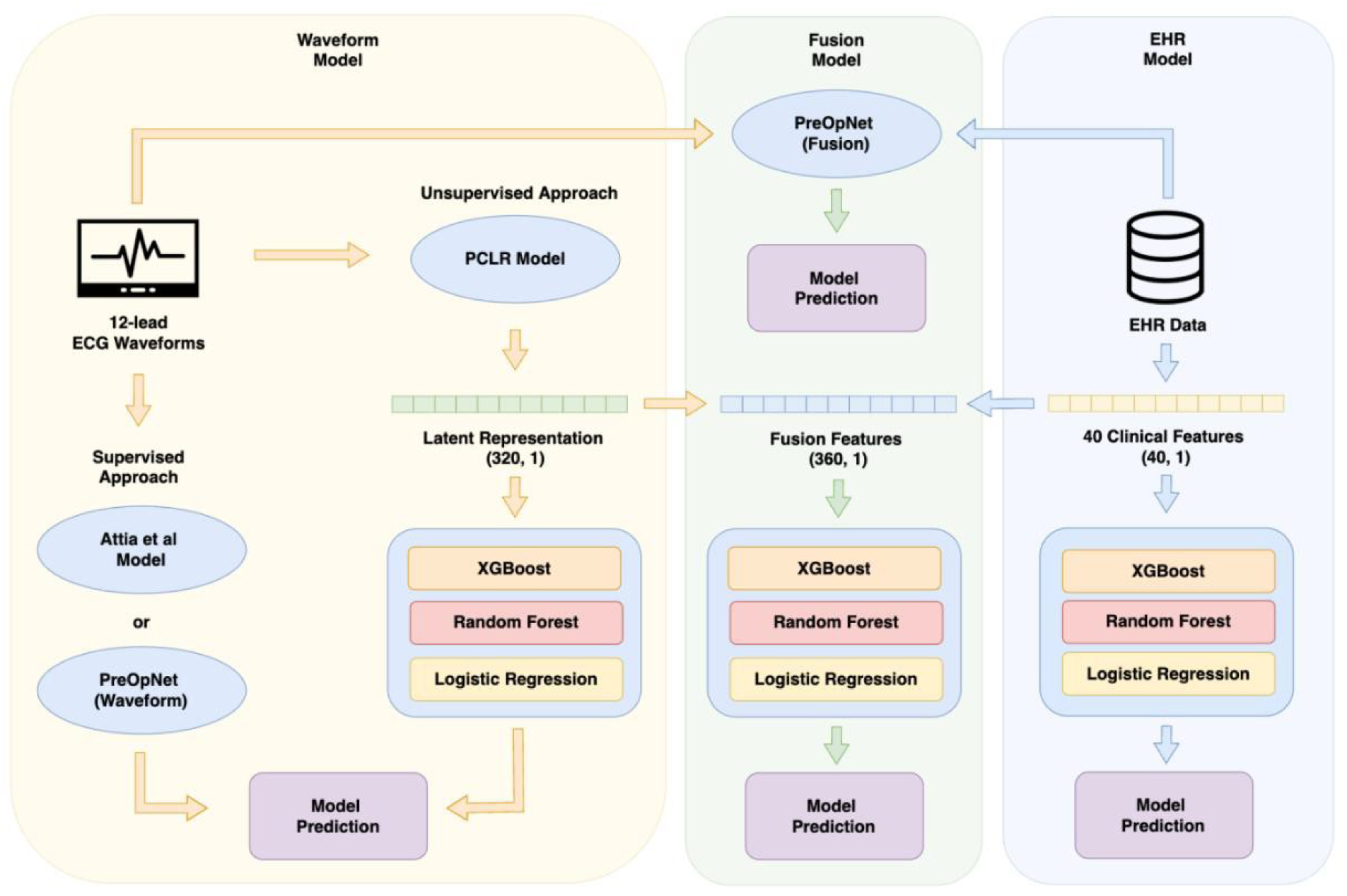
**Schematic overview of modeling approach**

The waveform models utilized raw 12-lead ECG waveforms of dimensions 12 x 5000, where 12 represents the 12 leads and 5000 represents the data points sampled at 500 Hz over 10 seconds. This input format was identical to the shape of the ECG waveforms from our dataset, requiring minimal preprocessing to the waveforms.

Three CNN-based architectures from previous literature were explored: Attia et al., PreOpNet, and Patient Contrastive Learning of Representations (PCLR) ^29–31^. CNN models were selected due to their capability to effectively capture temporal and spatial patterns within repetitive localized patterns such as the QRS complex and T-wave. The model from Attia et al. demonstrated success in atrial fibrillation (AF) detection, while PreOpNet showed the potential of CNNs in predicting post-procedural mortality. Both models are supervised, end-to-end approaches that directly generate final predictions from raw ECG input ^29,30^.

While these CNN models served as benchmarks, PCLR was our primary model for waveform analysis due to its unsupervised nature ^31^. Unsupervised approaches are well suited for situations where labeled data is scarce, as in the case with SCM due to SCM’s underdiagnosed nature and small number of SCM samples in our dataset. PCLR was pre-trained on a large dataset of 3.2 million unlabeled 12-lead ECGs from Massachusetts General Hospital (MGH) and Brigham and Women’s Hospital (BWH), producing a 320-dimensional latent representation of the ECG signal. To generate final predictions from these 320-dimensional representations, we applied three machine learning classifiers: XGBoost, Random Forest, and Logistic Regression^32,33^_._

The fusion models combined ECG waveforms with the 40 clinical features identified in the “Feature Extraction” section. Since current diagnosis of SCM relies on a combination of change in ECG patterns and various clinical indicators such as laboratory measurements, this fusion approach can leverage both data modalities to potentially enhance performance. Two CNN-based models, PreOpNet and PCLR, were explored due to their ability to integrate clinical features with ECG waveforms. The Attia et al model was excluded from the fusion model analysis as it only accepts ECG waveforms as input and is unable to integrate clinical features. In PreOpNet, ECG-derived features were concatenated with clinical features at the final fully connected layer before classification. For PCLR, the 320-dimensional ECG representation was concatenated with the 40 clinical features, resulting in a 360-dimensional input vector. This vector was then classified using the same three traditional machine learning classifiers, XGBoost, Random Forest, and Logistic Regression, to ensure consistency with the waveform model approach. To serve as a comparison to the waveform and fusion models, models were developed only utilizing clinical features. Machine learning approaches such as XGBoost, Random Forest, and Logistic Regression were explored, as these models are well suited for structured, tabular data.

### Model Development

To ensure robust hyperparameter optimization (HPO) and prevent data leakage, nested cross-validation (CV) with 5 outer loops and 3 inner loops was implemented for all models. For each iteration of the inner loop, a grid search was performed to identify the optimal hyperparameters using the area under the precision-recall curve (AUPRC) as the evaluation metric. The outer loop then evaluated model performance on unseen test data, ensuring a more robust estimate of model generalizability. The full list of hyperparameters explored and the optimal hyperparameters selected for the models is shown in Supplementary Table 3.

### Model Evaluation

The primary evaluation metric for this study was the area under the precision-recall curve (AUPRC) considering the severe class imbalance in our dataset. AUPRC is well suited for imbalanced datasets due to its emphasis on model performance on the minority class. While area under the receiver operating characteristic curve (AUROC) was also evaluated, it can be misleading in imbalanced datasets as the large number of true negatives (TNs) can inflate AUROC even for models with poor performance.

For threshold-dependent metrics such as sensitivity, specificity, F1 score, precision, positive predictive value (PPV), negative predictive value (NPV), and odds ratio (OR), the optimal threshold was determined based on the highest F1 score. Thresholds ranging from 0.01 to 0.99 were tested in increments of 0.01 to ensure sufficient granularity in thresholds evaluated. We additionally evaluated the Brier Score, a calibration metric that measures the mean squared difference between the actual labels and predicted probabilities, with lower Brier scores indicating better calibration.

### Statistical Analysis

95% confidence intervals (CIs) for all metrics were computed using bootstrapping with 10,000 resamples. To compare model performance and determine statistical significance, a permutation test with 10,000 resamples was conducted using a p-value threshold of 0.05. For comparing patient characteristics, z-test for proportions was used to determine statistical significance with 𝛼 = 0.05.

### Model Explainability

SHAP (SHapley Additive exPlanations) values were used for feature importance analysis in the EHR model ^34^. Strumbelj approximation, a modified SHAP approximation employed for the sake of computational tractability, was implemented for the fusion model to analyze the relative importance of ECG waveforms compared to the clinical features ^35^. This approach allowed for a detailed analysis of the contributions of each clinical feature and ECG to the final predictions and determining features of highest importance.

## Results

### Patient Characteristics

Among the 71,479 total patients in our dataset, 349 (0.48%) were identified as SCM patients while 4,507 (6.31%) were identified as AMI patients. Patient characteristics for all three cohorts are reported in Table 1. The SCM cohort had a mean age of 64.82 and a significantly higher proportion of females (81.1%) compared to non-SCM (45.6%, p < 0.001) and AMI (37.7%, p < 0.001). This finding aligns with previous literature that SCM predominantly affects older women. Comorbid congestive heart failure was significantly more common in SCM patients (49.7%) compared to non-SCM (26.8%, p < 0.001), but no significant difference was observed compared to AMI (50.3%, p = 0.83). The relatively high prevalence of comorbidities across all cohorts is expected, as all patients in our dataset were admitted to the hospital which elevates the likelihood of multiple coexisting medical conditions and chronic comorbidities.

**Table 1.**
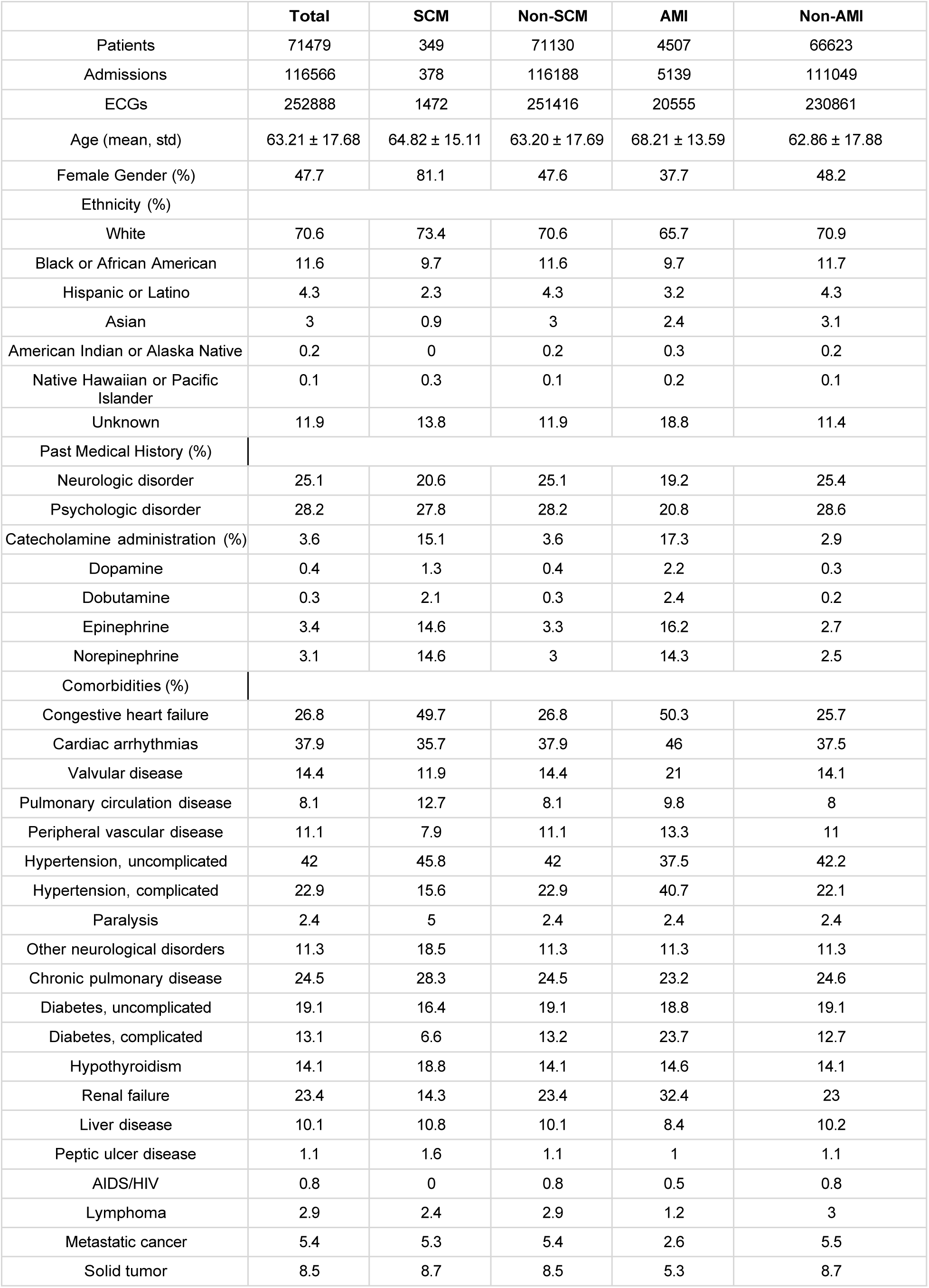
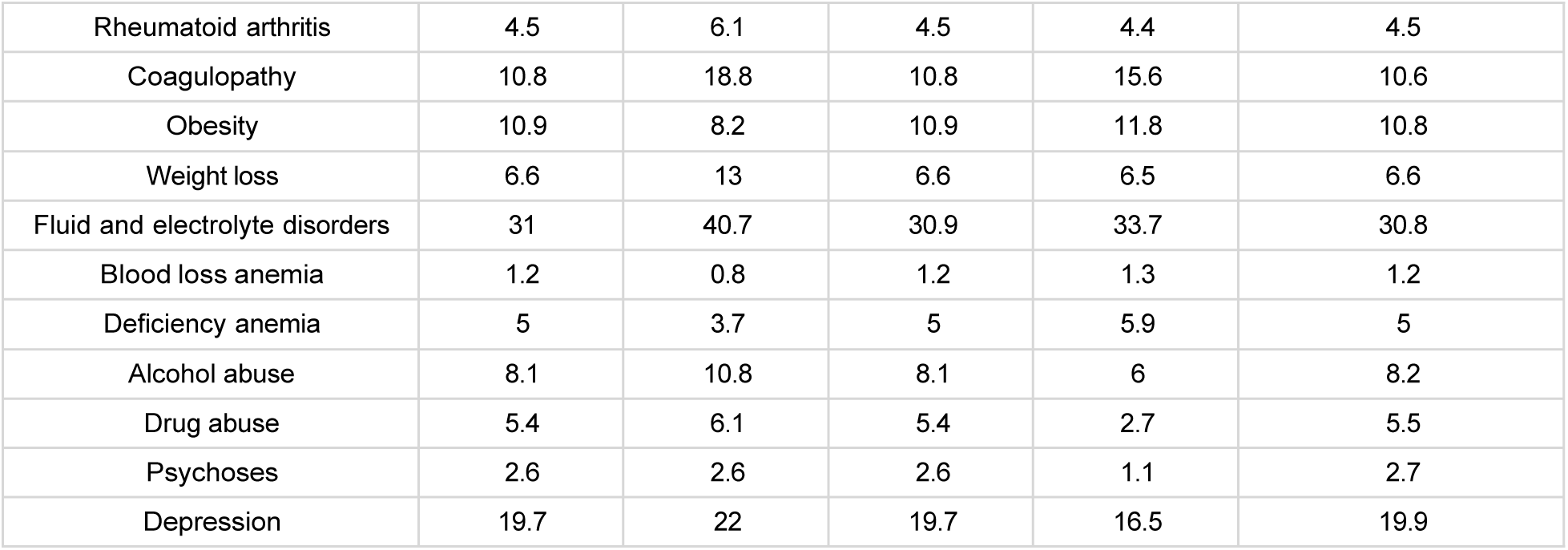
Patient Characteristics.

### Model Performance

Model performance for SCM vs. Non-SCM and SCM vs. AMI is shown in Table 2 and 3. Among all models, the fusion model using a PCLR + XGBoost classifier demonstrated the highest performance of AUPRC of 0.191 (0.146-0.243) for SCM vs. Non-SCM and 0.430 (0.371-0.488) for SCM vs. AMI, which outperformed the waveform and EHR models. These results are notable given the extremely imbalanced nature of our dataset, where baseline AUPRC values were 0.0048 for SCM vs. Non-SCM and 0.0625 for SCM vs. AMI. Baseline AUPRC values are the proportion of positive samples in the dataset (prevalence).

**Table 2.** Model Performance for SCM vs. Non-SCM.

| Model | Approach | AUROC | AUPRC | Brier Score | Sensitivity | Specificity | F1 Score | PPV | NPV | OR |
| --- | --- | --- | --- | --- | --- | --- | --- | --- | --- | --- |
| Waveform Model | Attia et al | 0.740<br>[0.709 - 0.768] | 0.025<br>[0.015 - 0.042] | 0.004<br>[0.003 - 0.004] | 0.076<br>[0.048 - 0.107] | 0.995<br>[0.994 - 0.995] | 0.061<br>[0.039 - 0.085] | 0.051<br>[0.032 - 0.072] | 0.997<br>[0.996 - 0.997] | 15.386<br>[9.303 – 22.442] |
|  | PreOpNet | 0.797<br>[0.715 - 0.865] | 0.064<br>[0.024 - 0.150] | 0.003<br>[0.002 - 0.004] | 0.146<br>[0.058 - 0.250] | 0.998<br>[0.997 - 0.999] | 0.161<br>[0.070 - 0.265] | 0.179<br>[0.073 - 0.303] | 0.997<br>[0.997 - 0.998] | 70.806<br>[28.671 – 134.947] |
|  | PCLR + XGBoost | 0.866<br>[0.846 - 0.887] | 0.089<br>[0.062 - 0.125] | 0.003<br>[0.003 - 0.004] | 0.160<br>[0.119 - 0.205] | 0.997<br>[0.997 - 0.998] | 0.169<br>[0.129 - 0.213] | 0.180<br>[0.135 - 0.229] | 0.997<br>[0.997 - 0.997] | 59.248<br>[43.240 – 79.728] |
|  | PCLR + Random Forest | 0.807<br>[0.780 - 0.833] | 0.045<br>[0.031 - 0.067] | 0.004<br>[0.003 - 0.004] | 0.135<br>[0.096 - 0.174] | 0.995<br>[0.994 - 0.995] | 0.107<br>[0.077 - 0.135] | 0.088<br>[0.062 - 0.114] | 0.997<br>[0.996 - 0.997] | 28.274<br>[19.657 – 37.623] |
|  | PCLR + Logistic Regression | 0.849<br>[0.825 - 0.873] | 0.092<br>[0.065 - 0.132] | 0.003<br>[0.003 - 0.004] | 0.219<br>[0.173 - 0.266] | 0.995<br>[0.995 - 0.996] | 0.172<br>[0.135 - 0.210] | 0.142<br>[0.111 - 0.174] | 0.997<br>[0.997 - 0.998] | 50.089<br>[38.197 – 64.165] |
| Fusion Model | PreOpNet | 0.813<br>[0.749 - 0.872] | 0.104<br>[0.045 - 0.199] | 0.004<br>[0.003 - 0.005] | 0.217<br>[0.111 - 0.328] | 0.998<br>[0.997 - 0.999] | 0.248<br>[0.130 - 0.360] | 0.289<br>[0.146 - 0.436] | 0.997<br>[0.996 - 0.998] | 98.388<br>[51.311 – 166.371] |
|  | PCLR + XGBoost | 0.918<br>[0.899 - 0.934] | 0.191<br>[0.146 - 0.243] | 0.003<br>[0.003 - 0.004] | 0.340<br>[0.283 - 0.395] | 0.996<br>[0.995 - 0.996] | 0.265<br>[0.221 - 0.309] | 0.217<br>[0.178 - 0.256] | 0.998<br>[0.997 - 0.998] | 91.034<br>[71.782 – 114.626] |
|  | PCLR + Random Forest | 0.868<br>[0.843 - 0.891] | 0.086<br>[0.062 - 0.124] | 0.003<br>[0.003 - 0.004] | 0.201<br>[0.157 - 0.255] | 0.996<br>[0.996 - 0.997] | 0.176<br>[0.136 - 0.220] | 0.156<br>[0.121 - 0.198] | 0.997<br>[0.997 - 0.997] | 54.159<br>[40.841 – 71.862] |
|  | PCLR + Logistic Regression | 0.906<br>[0.885 - 0.925] | 0.159<br>[0.118 - 0.204] | 0.003<br>[0.003 - 0.004] | 0.181<br>[0.137 - 0.225] | 0.999<br>[0.998 - 0.999] | 0.235<br>[0.182 - 0.284] | 0.338<br>[0.264 - 0.411] | 0.997<br>[0.997 - 0.997] | 114.313<br>[86.763 – 145.378] |
| EHR Model | XGBoost | 0.866<br>[0.843 - 0.890] | 0.078<br>[0.055 - 0.109] | 0.003<br>[0.003 - 0.004] | 0.236<br>[0.185 - 0.287] | 0.993<br>[0.993 - 0.994] | 0.154<br>[0.119 - 0.186] | 0.115<br>[0.087 - 0.142] | 0.997<br>[0.997 - 0.998] | 41.416<br>[31.355 – 52.519] |
|  | Random Forest | 0.884<br>[0.862 - 0.903] | 0.054<br>[0.041 - 0.076] | 0.003<br>[0.003 - 0.004] | 0.250<br>[0.201 - 0.298] | 0.989<br>[0.988 - 0.990] | 0.118<br>[0.093 - 0.141] | 0.077<br>[0.060 - 0.094] | 0.997<br>[0.997 - 0.998] | 28.205<br>[21.508 – 35.912] |
|  | Logistic Regression | 0.866<br>[0.844 - 0.886] | 0.040<br>[0.032 - 0.052] | 0.004<br>[0.003 - 0.004] | 0.128<br>[0.093 - 0.166] | 0.995<br>[0.994 - 0.995] | 0.098<br>[0.071 - 0.125] | 0.079<br>[0.056 - 0.101] | 0.997<br>[0.996 - 0.997] | 24.959<br>[17.760 – 33.170] |

For the waveform models, the PCLR + XGBoost classifier also outperformed other end-to-end CNN-based architectures, such as Attia et al. and PreOpNet, as well as other classifiers combinations with PCLR. The PCLR + XGBoost model achieved an AUPRC of 0.089 (0.062-0.125) for SCM vs. Non-SCM and 0.309 (0.257-0.374) for SCM vs. AMI.

The fusion model outperformed both the waveform model (p < 0.001) and the EHR model (p < 0.001) for both SCM vs. Non-SCM and SCM vs. AMI. There was no statistically significant difference in performance between the waveform model and the EHR model for both tasks.

A visual comparison of the best performing models from each category (waveform, fusion, EHR model) is shown in Figure 3.

**Figure 3.**
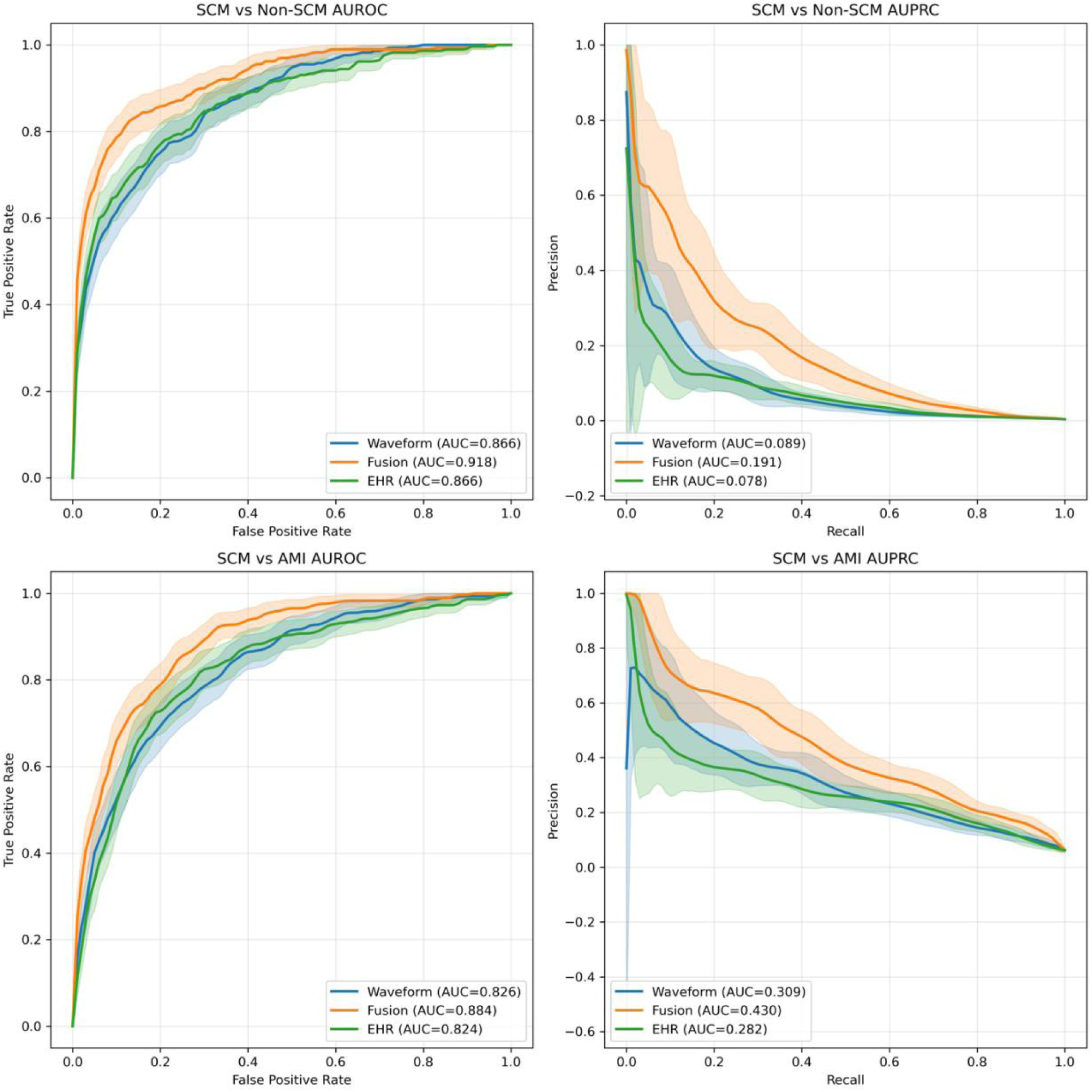
**Performance of Waveform, Fusion, and EHR Models**

### Model Explainability

Feature importance was analyzed using two methods: SHAP values for the best performing EHR model (XGBoost) and the Strumbelj approximation, a modified SHAP approach, for the best performing fusion model (PCLR + XGBoost).

Figure 4 shows the results for SHAP analysis for the XGBoost classifier on clinical features. Among the 40 clinical features, female gender, age, and comorbidities such as congestive heart failure (CHF), hypertension with complications, and diabetes with complications were identified as the most significant features in predicting SCM.

**Figure 4.**
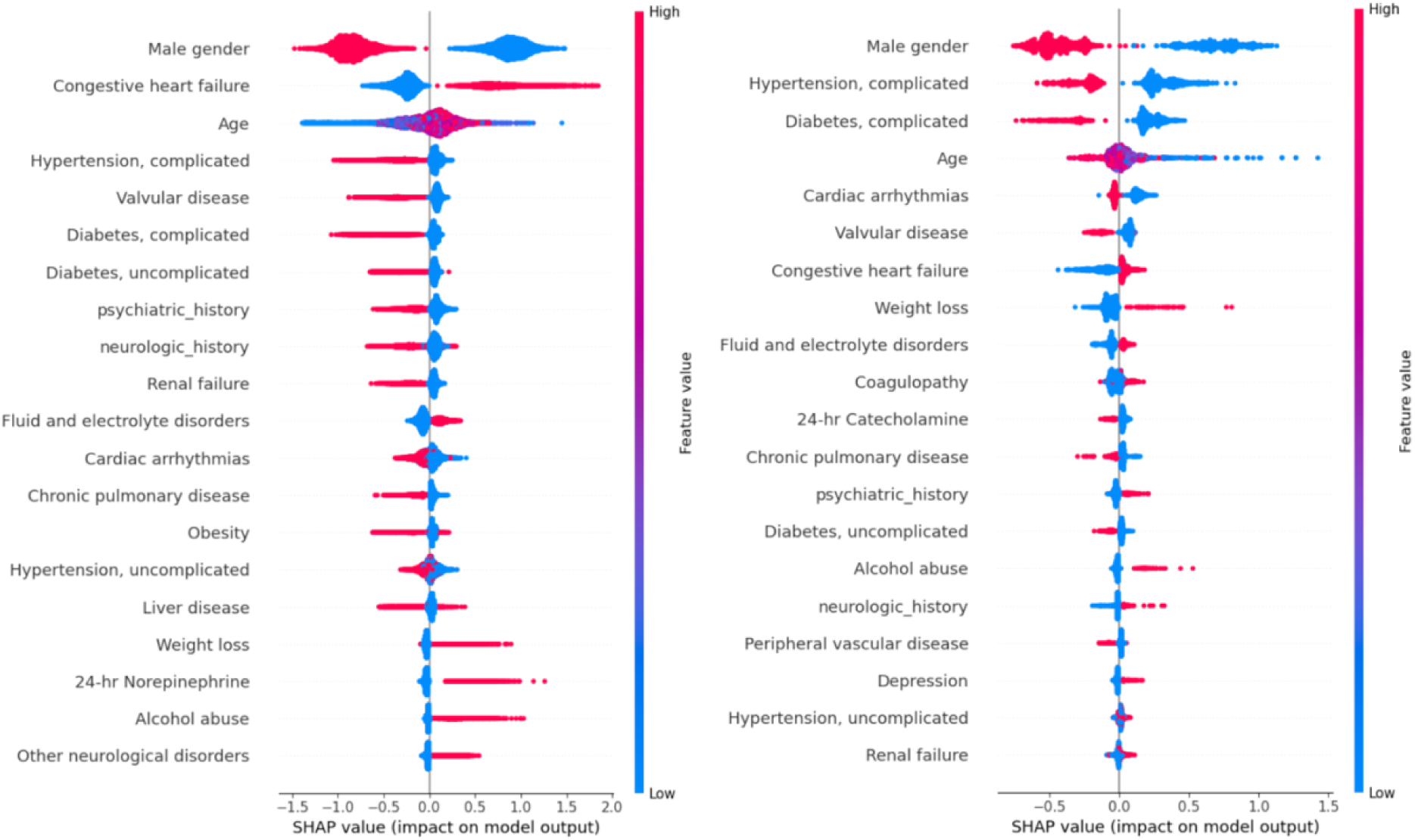
**SHAP Analysis on XGBoost approach for the EHR model. Left plot is for SCM vs non-SCM and right plot is for SCM vs AMI.**

Figure 5 shows the results of the Strumbelj approximation applied to the fusion model. This analysis helps us understand the relative importance of ECG features compared to individual clinical features. ECG features generated by the PCLR model were identified as the most critical features for both Tasks A and B. Clinical features such as gender, age, and comorbidities such as congestive heart failure (CHF), hypertension with complications, and diabetes with complications remained influential for model prediction.

**Figure 5.**
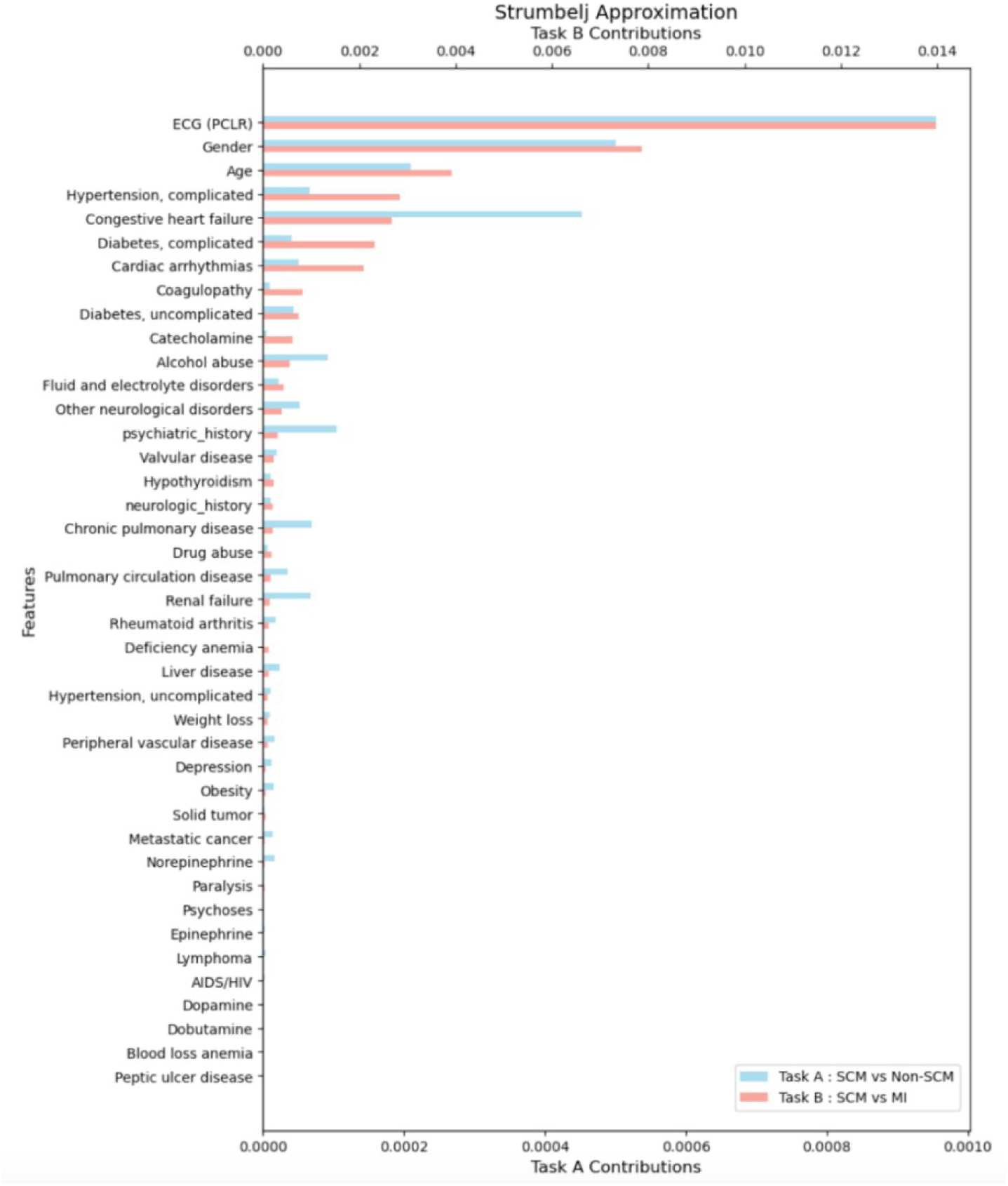
**Feature Importance Analysis for the Fusion Model**

## Discussion

### Key Results

Our findings indicate that raw ECG signals contain rich diagnostic information for identifying SCM. The superior performance of the fusion model compared to the waveform and EHR models suggests that ECG and clinical features are additive in determining predictive accuracy. Feature importance analysis aligns with previously reported risk factors for SCM. ^1,36^

### Analysis of previous literature

Previous studies of SCM have focused on echocardiography, cardiac magnetic resonance imaging, or traditional ECG analysis ^21,22,37–39^. Limitations of this prior research include small sample sizes (reducing generalizability) and artificially balanced class structures ranging from 1:1 to 1:3 for SCM versus control. This doesn’t reflect the true prevalence of SCM in real-world clinical settings, making it difficult to evaluate the clinical utility of these models. Additionally, most studies aimed to differentiate SCM from AMI, which does not address the clinical problem of identifying SCM among a wide range of non-SCM conditions in diverse clinical contexts.

Echocardiography, CMR, and brain MRI, are time-consuming and require specialized expertise for analysis ^14^. Implementation of these techniques may incur delay, and they are not suited for low-resource settings. Studies using traditional ECG features (e.g. ST level at the J-point or frequencies for ST-segment elevation, are extracted using methodologies that might be specific to individual devices or hospitals ^22^. This lack of standardization limits the generalizability of these studies to be used in an ECG machine-agnostic manner ^23,24^.

### Strengths

This study introduces a novel approach of utilizing raw 12-lead ECG waveforms to differentiate SCM. To the best of our knowledge this is the first study to utilize this approach, which allows for rapid screening compared to methods used in previous studies. Leveraging 12-lead ECG analysis could streamline the screening workflow, especially in low-resource settings, to raise suspicion for SCM and refer patients to centers with more advanced diagnostic capabilities. The waveform model’s strong performance shows the potential of ECG waveforms to become a paradigm shift in SCM screening and diagnosis. This approach also allows for an ECG machine-agnostic solution that does not depend on embedded ECG parameter extraction processes. It also increases the likelihood that SCM patients receive timely and accurate diagnosis and therapeutic intervention, preventing misclassification as AMI or other acute coronary syndromes and reducing risk of exposure to potentially harmful and costly testing such as coronary angiography.

This study analyzes one of the largest samples among similar studies, including a total of 71,479 patients with 349 SCM and 4,507 AMI patients. This large sample size allows for training and evaluating our model on a more diverse population, increasing robustness and generalizability. Also, by maintaining a class imbalance of 0.3% for SCM vs. non-SCM and 6.3% for SCM vs. AMI, this study reflects the real-world prevalence of SCM, allowing for a more realistic model performance evaluation in clinical settings. Lastly, in contrast to previous studies that focused exclusively on SCM vs. AMI, this work considers the SCM vs. non-SCM distinction, which points to utility in a broader clinical context.

### Limitations

This work has several limitations. First, we did not perform external validation to evaluate model robustness in other clinical environments. We searched other publicly available 12-lead ECG datasets, such as the China Physiological Signal Challenge 2018 (CPSC) dataset, the Georgia 12-Lead ECG Challenge Database, and the PTB-XL dataset, none of which had labels for SCM ^40–42^. Second, the MIMIC-IV dataset does not contain key features linked to SCM, such as stressor exposure and cardiac biomarkers. SCM stressors might include recent physical stress, such as major surgery, trauma, stroke or seizure, or recent emotional stress, such as loss of a family member or occupation ^1,2^. The identification of a recent stressor is a cardinal diagnostic feature of SCM, however such information was not available in the MIMIC-IV database. For serum cardiac biomarkers like troponin or pro-BNP, data was missing in over 70% of our cohort, preventing rigorous integration into the model.

A third limitation arises from the use of ICD-9/10 codes as ground truth labels. These codes are assigned to entire hospital admissions, making it difficult to identify the exact timing of SCM onset. Therefore, if SCM developed midway through admission, early ECG measurements might not reflect characteristics of SCM. In addition, a small number of patients (n = 62) carried dual diagnoses of SCM and AMI. While these patients were excluded from the SCM vs. AMI task to avoid label ambiguity, they were retained in the SCM vs. Non-SCM analysis. Given their small proportion (< 0.1% of the cohort), we do not expect their inclusion to significantly affect model performance.

Finally, the severe class imbalance, especially in the SCM vs. non-SCM task, posed challenges in preventing overfitting. While data augmentation techniques such as Synthetic Minority Oversampling Technique (SMOTE) are commonly used in other domains, there is a lack of widely validated augmentation methods for time-series data such as ECGs ^43,44^. This class imbalance was addressed through a rigorous nested cross-validation framework and a pre-trained, unsupervised approach (PCLR) to learn generalizable ECG representations before fine-tuning on the imbalanced dataset, achieving superior performance compared to end-to-end approaches like Attia et al. and PreOpNet. However, this unsupervised approach prevented us from applying model explainability techniques such as Gradient-weighted Class Activation Mapping (GradCAM) directly on the ECG waveforms, limiting interpretability of the model ^45^.

### Future Directions

Further establishing the validity of our approach can be accomplished in prospective studies that meticulously document stressor exposure and other features. Also, external validation on other large ECG datasets with SCM labels, once available, will be critical to assess generalizability. More advanced modeling approaches, such vision transformers applied to ECGs, might further enhance model performance. Finally, while this study focuses on rapid and resource-efficient diagnosis, exploring comprehensive fusion models that integrate additional modalities such as echocardiography or CMR could provide even greater diagnostic accuracy in well-resourced environments.

## Data Availability

The data that support the findings of this study are publicly available from the PhysioNet repository as part of the Medical Information Mart for Intensive Care IV (MIMIC-IV) database, including the MIMIC-IV hosp, MIMIC-IV-ECG, and MIMIC-IV-Note modules. Access to the dataset is provided under a data use agreement and requires completion of required training and credentialing. The MIMIC-IV database is available at: https://physionet.org/content/mimiciv/

## Acknowledgments

The authors thank Tianyou Liu, an undergraduate student at Johns Hopkins University, for his assistance with the literature search that supported the background and contextualization of this study.

---

**Table 3.** Model Performance for SCM vs. AMI.

| Model | Approach | AUROC | AUPRC | Brier Score | Sensitivity | Specificity | F1 Score | PPV | NPV | OR |
| --- | --- | --- | --- | --- | --- | --- | --- | --- | --- | --- |
| Waveform Model | Attia et al | 0.810<br>[0.782 - 0.835] | 0.287<br>[0.244 - 0.348] | 0.051<br>[0.046 - 0.056] | 0.389<br>[0.333 - 0.447] | 0.951<br>[0.945 - 0.958] | 0.367<br>[0.321 - 0.418] | 0.348<br>[0.298 - 0.400] | 0.959<br>[0.953 - 0.965] | 8.474<br>[6.925 – 10.551] |
|  | PreOpNet | 0.698<br>[0.627 - 0.763] | 0.292<br>[0.199 - 0.397] | 0.077<br>[0.062 - 0.092] | 0.350<br>[0.247 - 0.457] | 0.935<br>[0.919 - 0.950] | 0.339<br>[0.237 - 0.431] | 0.329<br>[0.222 - 0.427] | 0.940<br>[0.925 - 0.955] | 5.518<br>[3.442 – 8.274] |
|  | PCLR + XGBoost | 0.826<br>[0.801 - 0.850] | 0.309<br>[0.257 - 0.374] | 0.056<br>[0.050 - 0.063] | 0.424<br>[0.367 - 0.481] | 0.938<br>[0.931 - 0.945] | 0.361<br>[0.315 - 0.405] | 0.314<br>[0.269 - 0.360] | 0.961<br>[0.955 - 0.966] | 7.997<br>[6.444 – 9.874] |
|  | PCLR + Random Forest | 0.776<br>[0.748 - 0.805] | 0.220<br>[0.182 - 0.265] | 0.054<br>[0.049 - 0.059] | 0.354<br>[0.298 - 0.408] | 0.942<br>[0.935 - 0.949] | 0.319<br>[0.272 - 0.362] | 0.291<br>[0.243 - 0.337] | 0.956<br>[0.950 - 0.962] | 6.654<br>[5.338 – 8.157] |
|  | PCLR + Logistic Regression | 0.814<br>[0.788 - 0.838] | 0.277<br>[0.230 - 0.333] | 0.053<br>[0.048 - 0.058] | 0.413<br>[0.359 - 0.476] | 0.941<br>[0.934 - 0.947] | 0.358<br>[0.312 - 0.406] | 0.316<br>[0.271 - 0.363] | 0.960<br>[0.955 - 0.966] | 7.929<br>[6.447 – 9.912] |
| Fusion Model | PreOpNet | 0.740<br>[0.670 - 0.800] | 0.273<br>[0.177 - 0.386] | 0.066<br>[0.051 - 0.082] | 0.368<br>[0.254 - 0.479] | 0.943<br>[0.928 - 0.957] | 0.352<br>[0.245 - 0.451] | 0.338<br>[0.224 - 0.443] | 0.949<br>[0.934 - 0.964] | 6.670<br>[4.050 – 10.233] |
|  | PCLR + XGBoost | 0.884<br>[0.863 - 0.903] | 0.430<br>[0.371 - 0.488] | 0.051<br>[0.045 - 0.057] | 0.427<br>[0.371 - 0.489] | 0.966<br>[0.961 - 0.971] | 0.441<br>[0.387 - 0.493] | 0.456<br>[0.398 - 0.511] | 0.962<br>[0.956 - 0.968] | 11.982<br>[9.852 – 14.970] |
|  | PCLR + Random Forest | 0.830<br>[0.806 - 0.856] | 0.318<br>[0.264 - 0.376] | 0.050<br>[0.045 - 0.055] | 0.448<br>[0.388 - 0.505] | 0.944<br>[0.937 - 0.950] | 0.392<br>[0.341 - 0.435] | 0.349<br>[0.299 - 0.394] | 0.963<br>[0.957 - 0.968] | 9.297<br>[7.514 – 11.438] |
|  | PCLR + Logistic Regression | 0.848<br>[0.824 - 0.871] | 0.339<br>[0.287 - 0.402] | 0.051<br>[0.046 - 0.056] | 0.431<br>[0.373 - 0.492] | 0.951<br>[0.945 - 0.958] | 0.399<br>[0.346 - 0.449] | 0.371<br>[0.321 - 0.426] | 0.962<br>[0.956 - 0.967] | 9.680<br>[7.863 – 11.969] |
| EHR Model | XGBoost | 0.824<br>[0.797 - 0.848] | 0.282<br>[0.240 - 0.337] | 0.051<br>[0.046 - 0.056] | 0.531<br>[0.473 - 0.588] | 0.896<br>[0.888 - 0.905] | 0.344<br>[0.304 - 0.381] | 0.254<br>[0.220 - 0.287] | 0.966<br>[0.961 - 0.971] | 7.546<br>[6.078 – 9.430] |
|  | Random Forest | 0.820<br>[0.793 - 0.844] | 0.282<br>[0.235 - 0.333] | 0.051<br>[0.047 - 0.056] | 0.514<br>[0.455 - 0.569] | 0.907<br>[0.898 - 0.916] | 0.353<br>[0.311 - 0.392] | 0.269<br>[0.234 - 0.304] | 0.966<br>[0.960 - 0.971] | 7.788<br>[6.292 – 9.507] |
|  | Logistic Regression | 0.810<br>[0.783 - 0.836] | 0.265<br>[0.219 - 0.318] | 0.052<br>[0.048 - 0.057] | 0.552<br>[0.493 - 0.612] | 0.881<br>[0.871 - 0.890] | 0.331<br>[0.291 - 0.370] | 0.236<br>[0.205 - 0.269] | 0.967<br>[0.961 - 0.973] | 7.198<br>[5.741 – 9.024] |

**Supplementary Table 1.** ICD 9/10 codes used for SCM and AMI.

| Condition | ICD Version | Code Pattern | Codes | Codes not included |
| --- | --- | --- | --- | --- |
| SCM | 9 | 429.XX | 429.83 | - |
|  | 10 | I51.XX | I51.81 | - |
| AMI | 9 | 410.XX | 410.00, 410.01, 410.02, 410.10, 410.11, 410.12, 410.20, 410.21, 410.22, 410.30, 410.31, 410.32, 410.40, 410.41, 410.42, 410.50, 410.51, 410.52, 410.80, 410.81, 410.82, 410.90, 410.91, 410.92 | 410.60, 410.61, 410.62 (True posterior wall infarction)<br>410.70, 410.71, 410.72 (Subendocardial infarction) |
|  | 10 | I21.X<br>X<br>I21.A<br>X<br>I22.X<br>I23.X | I21.01, I21.02, I21.09, I21.10, I21.11, I21.19, I21.20, I21.21, I21.29, I21.3, I21.4, I21.9, I21.A, I21.A1, I21.A9, I22.0, I22.1, I22.2, I22.8, I22.9, I23, I23.0, I23.1, I23.2, I23.3, I23.4, I23.5, I23.6, I23.7, I23.8 | - |

**Supplementary Table 2.** Feature Extraction.

| Category | Features | Notes |
| --- | --- | --- |
| Demographics | Age, Gender |  |
| Comorbidities | Congestive Heart Failure, Coronary Artery Disease, Valvular Disease, Peripheral Vascular Disease, Pulmonary Vascular Disease, Hypertension (Uncomplicated), Hypertension (Complicated), Paraplegia, Other Neurological Disorders, Chronic Pulmonary Disease, Diabetes (Uncomplicated), Diabetes (Complicated), Hypothyroidism, Renal Failure, Liver Disease, Peptic Ulcer Disease, Acquired Immunodeficiency Syndrome (AIDS), Lymphoma, Metastatic Cancer, Solid Tumor without Metastasis, Rheumatoid Arthritis or Connective Tissue Disease, Functional or Cognitive Impairment, Blood Anemia, Drug/Alcohol-Induced Anemia, Alcohol Abuse, Drug Abuse, Psychiatric Disorders, Depression | Elixhauser Comorbidity Index |
| Medical History | Psychiatric History | Keyword matching:<br>'depression', 'anxiety', 'schizophrenia', 'bipolar', 'ptsd', 'ocd', 'adhd', 'panic', 'eating disorder', 'anorexia', 'bulimia', 'borderline personality', 'psychosis', 'substance abuse', 'addiction', 'alcoholism', 'insomnia', 'dysthymia', 'major depressive disorder', 'cyclothymia', 'social anxiety', 'generalized anxiety', 'adjustment disorder', 'phobia' |
|  | Neurologic History | Keyword matching:<br>'migraine', 'stroke', 'epilepsy', 'parkinson', 'dementia', 'alzheimer', 'vascular dementia', 'multiple sclerosis', 'als', 'amyotrophic lateral sclerosis', 'huntington', 'neuropathy', 'traumatic brain injury', 'tbi', 'cerebral palsy', 'encephalopathy', 'meningitis', 'myasthenia gravis', 'guillain-barré', 'neuralgia', 'trigeminal neuralgia', 'concussion', 'brain tumor', 'bell's palsy', 'hydrocephalus' |
| Catecholamine Administration | Catecholamine, Dopamine, Epinephrine, Norepinephrine, Dobutamine | Excluded Medications:<br>Lidocaine, bupivacaine, racepinephrine, epipen |

**Supplementary Table 3.** Hyperparameter grid.

| Model | Hyperparameter Grid |
| --- | --- |
| XGboost | 'n_estimators': [100, 200, 300],<br>'max_depth': [3, 5, 7],<br>'learning_rate': [0.01, 0.1, 0.2],<br>'subsample': [0.6, 0.8, 1.0],<br>'colsample_bytree': [0.6, 0.8, 1.0],<br>'min_child_weight': [1, 3, 5],<br>'gamma': [0, 0.1, 0.2], |
| Random Forest | 'n_estimators': [50, 100, 200],<br>'max_depth': [None, 10, 20],<br>'min_samples_split': [2, 5, 10],<br>'min_samples_leaf': [1, 2, 4],<br>'max_features': ['sqrt', 'log2',<br>None] |
| Logistic Regression | 'C': [0.01, 0.1, 1, 10, 100],<br>'penalty': ['l1',<br>'l2'], 'solver':<br>['liblinear'] |

